# Assessing the Profile of Unvaccinated COVID-19 Individuals in African American and Latinx Communities in Eastern Pennsylvania

**DOI:** 10.1101/2022.02.11.22270504

**Authors:** Kenya M. Colvin, Kennedy S. Camara, Latasha S. Adams, Adline P. Sarpong, Danielle G. Fuller, Sadie E. Peck, Anthony S. Ramos, Ariana L. Acevedo, Meless A. Badume, Shae-lyn A. Briggs, Tiffany N. Chukwurah, Zanett Davila-Gutierrez, James A. Ewing, Jemimah O. Frempong, Amirah A. Garrett, Steven J. Grampp, Jahasia W. Gillespie, Emmanuel J. Herrera, Emis J. Maddox, John C. Pelaez, Olivia L. Quartey, Fanny Rodriguez, Luis A. Vasquez, Brian J. Piper, Swathi Gowtham

**Author notes:** authors contributed equally and were listed in alphabetical order (Ariana L. Acevedo, Meless A. Badume, Shae-lyn A. Briggs, Tiffany N. Chukwurah, Zanett Davila-Gutierrez, James A. Ewing, Jemimah O. Frempong, Amirah A. Garrett, Steven J. Grampp, Jahasia W. Gillespie, Emmanuel J. Herrera, Emis J. Maddox, John C. Pelaez, Olivia L. Quartey, Fanny Rodriguez, Luis A. Vasquez). Corresponding Author: Swathi Gowtham, MD, Geisinger Commonwealth School of Medicine, Scranton, PA, 18509, Pediatric Infectious Diseases, Department of Pediatrics, Geisinger Medical Center, Danville, PA 17822.

## Abstract

**Background:** Throughout US history, chronic and infectious diseases have severely impacted minority communities due to lack of accessibility to quality healthcare, accurate information, and underlying racism. These fault lines in the care of minority communities in the US have been further exposed by the rise of COVID-19 pandemic. This study examined the factors associated with COVID-19 vaccine hesitancy among African American and Latinx communities in Eastern Pennsylvania (PA).

**Methods:** Survey data was collected in July 2021 in Philadelphia, Scranton, Wilkes-Barre, and Hazleton, PA. The 203 participants (38.7% Black, 27.5% Latinx) completed the 28-question survey of COVID-19 vaccination attitudes in either English or Spanish.

**Results:** Out of a total of 181 participants that met inclusion criteria of completed surveys, results indicate that 63.5% (n=115) were acceptant of the COVID-19 vaccine whereas the remainder 36.5% (n=66) were hesitant. Binary logistic regression results showed that age, concern for vaccine efficacy, race, knowledge on the vaccine, and belief that the COVID-19 virus is serious significantly influenced COVID vaccine hesitancy. Minorities were more likely to be hesitant toward vaccination (OR: 2.77, 95% CI: 1.13, 6.79) than non-Hispanic whites. Those who believed the COVID vaccine was ineffective (OR: 8.29, 95% CI: 3.78,18.2), and that the virus is not serious (OR: 8.28, 95% CI: 1.11, 61.8) showed the greatest odds of hesitancy.

**Conclusions:** Contributing factors of vaccine hesitancy in minority communities were age, concern for vaccine efficacy, and education. Understanding and addressing the barriers to COVID-19 vaccination in minority groups is essential to decreasing transmission and controlling this pandemic.

## Background

Health disparities are differences in disease prevention measures and health outcomes in socially disadvantaged communities [1]. Socially disadvantaged communities can include certain racial and ethnic groups, LGBTQ+ communities, rural versus urban communities, and low socioeconomic areas [1]. Racial and ethnic health disparities have been evident in healthcare in the US for many years. For example, African American and Latinx communities are more likely to suffer from cardiovascular diseases compared to non-Hispanic whites [2]. Researchers have attributed these disparities to several factors, including socioeconomic status, levels of education, structural racism, medical mistrust and lack of quality healthcare access [2, 3, 4]. Low socioeconomic status has negatively influenced health outcomes for socially disadvantaged communities. This association is attributed to individuals with a low socioeconomic status lacking health insurance or lacking access to quality healthcare.

This has been further evident in worse health outcomes in socially disadvantaged communities regarding COVID-19. Lower socioeconomic communities have higher prevalence rates of contracting COVID-19 compared to higher socioeconomic groups [5]. Geographical concentration of minorities in urban areas where many work in public service jobs may also contribute to the disproportionate, negative impact COVID-19 has had on minority communities across the US [6]. Urban settings are more likely to be densely populated and have limited capacity for social distancing [6]. African American and Hispanic/Latinx communities across the US have experienced higher rates of infection and death due to COVID-19 compared to non-Hispanic whites [7,8]. Higher mortality rates amongst these communities may be due to higher comorbidities, such as cardiovascular illnesses, diabetes, and lack of timely access to quality healthcare [2,6].

Despite the increased burden of disease, African American and Latinx communities displayed the highest levels of hesitancy for the COVID vaccine in the US [9]. Vaccine hesitancy has been a major, global public health concern for many years, and has gained increasing attention as the world races to end the coronavirus pandemic [7,10]. Vaccine hesitancy exists on a spectrum and is reluctance or complete refusal to receive a vaccine regardless of availability due to a complex entanglement of epidemiological factors [10]. Researchers have found vaccine hesitancy to be context-specific, meaning that individuals who are willing to take one vaccine may be reluctant or refuse to take other vaccines [8]. Although vaccine hesitancy is generally well documented, data on vaccine hesitancy in the context of the COVID-19 vaccine is limited and continually evolving as attitudes towards vaccination shift [11,12]. Several national studies predicting COVID-19 vaccine uptake in the US were published prior to the vaccines becoming available [13, 14]. One large, national survey-based study found vaccine hesitancy among African Americans to be 34% and 29% among Hispanics [10].

When COVID-19 vaccines became available, hesitancy among these two major ethnic groups remained high in part due to inequities in vaccine distribution across the US [7]. Since vaccine hesitancy is context-specific, recommended mitigation strategies often include engaging with local communities to collectively develop solutions that address their reluctance toward vaccination [7, 8]. Despite several studies being conducted at the national level [7,9,14], few studies have examined factors driving hesitancy among ethnic minorities by state and even fewer for the state of Pennsylvania (PA) [15, 16]. Our research aimed to identify factors influencing vaccine disparities in eastern PA. Gaining a deeper understanding of these underlying issues may lead to effective, community specific solutions that promote vaccine uptake. In the first year of the pandemic, African Americans and Latinx communities in PA faced a rate of COVID-19 infection (18% and 21% of cases respectively) greater than their percentage of the state’s population (11% and 7% of the population respectively) [17]. The dynamic nature of the COVID-19 pandemic requires continued monitoring of vaccination uptake rates as this directly impacts the ability to end the pandemic worldwide [7]. The recent surge in cases and hospitalization rates in PA and across the US demonstrate the urgent need to increase vaccination uptake in minority communities [18]. The objectives of this study were to outline the major influences on COVID-19 vaccine hesitancy among Latinx and African Americans in Eastern PA and promote vaccine acceptance.

## Methods

### Participants

Data for the participants was collected from a convenience sample in the Philadelphia, Hazleton, Scranton, and Wilkes-Barre areas of Eastern PA. The target population for this study were people ages 18 and older who lived in these areas that had access to information about the vaccine, with a focus on the underrepresented minority population. Among the participants, 198 (97.5%) individuals filled the survey out in English and 5 (2.5%) in Spanish. The ages of the participants ranged from 18 to over The sample population contained nearly equal amounts of men and women. Over half of the participants identified as African American or Latinx (Table 1).

**Table 1.** Demographics of participants in eastern-Pennsylvania completing a survey of COVID-19 vaccination attitudes. COVID-19 vaccine information sources included: my doctor, major news channels, the newspaper, social media, family members, or other. Information sources for the COVID-19 disease included: social media, local news, CDC, local pharmacy, medical professionals, family/friends, coworkers or other.

| Variable | Percentage of Respondents<br>n (%) | Total<br>(N) |
| --- | --- | --- |
| Age |  |  |
| Under 45 (18-45) | 146 (74.1) |  |
| Over 45 (45-65+) | 51 (25.9) | 197 |
| Gender |  |  |
| Male | 97 (49.2) |  |
| Female | 100 (50.8) | 197 |
| Education Level |  |  |
| Secondary | 116 (59.4) |  |
| Some High School (No Diploma) |  |  |
| High School Diploma or GED |  |  |
| Post-Secondary |  |  |
| Associate Degree | 79 (40.5) |  |
| Bachelor's Degree |  |  |
| Master's Degree |  |  |
| Doctorate's Degree |  |  |
| Professional Degree beyond Bachelor's |  | 195 |
| Race |  |  |
| Non-Hispanic White | 57 (29.4) |  |
| Minority | 139 (70.6) |  |
| Black or African American | 76 (38.7) |  |
| Hispanic, Latinx, or of Spanish Origin | 54 (27.5) |  |
| Asian | 9 (4.6) |  |
| American Indian or Alaska Native | 1 (0.51) |  |
| Middle Eastern or North African | 1 (0.51) |  |
| Native Hawaiian or other Pacific Islander | 0 (0) |  |
| Multiethnic | 7 (3.5) | 196 |
| Health Insurance |  |  |
| Yes | 165 (85.9) |  |
| No | 27 (14.0) | 192 |
| Affected by COVID-19 |  |  |
| Not Affected | 48 (25.0) |  |
| Affected | 144 (75.0) | 192 |
| "If you have not gotten the vaccine,<br>how safe do you think the vaccine is?" |  |  |
| Safe | 18 (25.0) |  |
| Not safe | 54 (75.0) | 72 |
| "I have concerns that the COVID-19 vaccine<br>is not effective." |  |  |
| Agree | 88 (48.1) |  |
| Disagree | 95 (51.9) | 183 |
| "If you do not want to get the vaccine,<br>which answer best describes your reason<br>for not getting the vaccine?" |  |  |
| Personal reasons | 30 (42.2) |  |
| Distrustful of |  |  |
| scientific community | 41 (57.7) | 71 |
| Concerned about getting COVID-19 again |  |  |
| Not worried | 19 (54.2) |  |
| Worried | 16 (45.7) | 35 |
| Paid sick leave |  |  |
| Yes | 124 (64.9) |  |
| No | 67(35.0) | 191 |
| History of COVID-19 |  |  |
| Yes | 35 (17.8) |  |
| No | 162 (82.1) | 197 |
| If no history of COVID-19, how worried are you about getting it? |  |  |
| Not worried | 77 (49.4) |  |
| Worried | 79 (50.6) | 156 |
| "How serious do you think the COVID-19 virus is?" |  |  |
| Serious | 182 (94.7) |  |
| Not serious | 10 (5.21) | 192 |
| Knowledgeable about the COVID-19 vaccine |  |  |
| Knowledgeable | 166 (89.2) |  |
| Not Knowledgeable | 20 (10.8) | 186 |
| "Where do you get most of your information about the vaccine?" |  |  |
| More than 3 sources | 17 (9.3) |  |
| Three or fewer sources | 166 (90.7) | 183 |
| Impact of COVID-19 Information Sources on the community |  |  |
| Positive | 99 (53.2) |  |
| Negative | 87 (46.8) | 186 |
| "Where do you obtain your information on COVID-19" |  |  |
| More than 3 sources | 17 (9.2) |  |
| Three or fewer sources | 167 (90.8) | 184 |
| Vaccine Hesitancy |  |  |
| Acceptant | 115 (63.5) |  |
| Like to get it as soon as possible | 6 (3.3) |  |
| I have already gotten it | 111 (61.3) |  |
| Hesitant | 66 (36.4) |  |
| Never get the vaccine | 26 (14.4) |  |
| Only if it is required by the state or employer | 8 (4.4) |  |
| Wait and see | 32 (17.7) | 181 |

### Procedures

This study was modeled after previously used surveys found on PubMed and the data was collected using SurveyMonkey. Members of the Center of Excellence summer program (COE) identified high traffic areas in the cities of interest. Personal protective equipment was worn throughout data collection in accordance with CDC guidelines to ensure the safety of the research team as well as the public. The study was conducted throughout July 2021 using a script to maintain consistency across sites. Investigators obtained verbal consent as well as verbal confirmation of participants being at least 18 years of age prior to survey completion. Educational materials were developed and distributed with information gathered about the vaccines in the form of fact sheets. Links to resources and different community organizations involved in vaccination efforts were also shared with participants.

The survey contained 28 questions organized into the following three categories: demographics, attitude towards COVID-19 disease, and attitude towards the COVID-19 vaccine (Table S1). We coded 19 of the 28 questions into dichotomous variables by grouping response choices for each category into two options. Open-ended response questions and questions that were unable to be dichotomized into two distinct groups are not presented here. All procedures were approved as exempt by the Institutional Review Board of Geisinger.

### Data Analysis

Participants who answered less than 10% of the survey were removed (N=3). Descriptive analysis was performed separately on each categorical variable to characterize the participants’ socio-demographics (Table 1). Multinomial logistic regression was employed to determine which factors had the greatest influence on vaccine hesitancy. To develop a regression model, 17 of the 28 questions were dichotomized (Table 1). For gender, there were three categories: male, female, and non-binary gender. The subset of participants who identified as non-binary gender (N=3) were not included in this analysis. Binary logistic regression was performed first to identify which factors had a statistically significant impact on vaccine hesitancy. The results were considered statistically significant if the p-value was less than 0.05. The significant factors were then incorporated into the multinomial logistics regression model. Missing values were removed for the descriptive analyses and both binary and multinomial regression. Multinomial logistic regression was used due to the methods effectiveness at analyzing multiple categorical variables. A forward regression was run using vaccine acceptance as the reference category for the dependent variable of vaccine hesitancy. The data analysis was conducted using IBM SPSS Statistics 28. Figures were developed using GraphPad Prism 9.3.1 for Windows.

## Results

Table 1 illustrates the demographics of the participants. Education level was dichotomized to secondary and post-secondary. Most participants (n=116, 59.7%) had secondary educational qualification (some high school to a high school diploma) while post-secondary (associates degree and beyond) accounted for the remaining two-fifths (n=78, 40.2%). The vast majority (n=165, 85.9%) of participants had health insurance although one-seventh (n=27, 14.1%) did not.

To determine which factors had the most influence on vaccine hesitancy, binary logistic regression was performed for each dichotomized variable in Table 1. The following variables had a statistically significant impact on vaccine hesitancy: age (p ≤ 0.001), concerned vaccine not effective (p ≤ 0.001), race (p ≤ 0.003), knowledgeable about the COVID-19 vaccine (p = 0.008), and how serious the COVID-19 virus is (p ≤ 0.020).

Supplemental Table 2 depicts the most influential factors associated with COVID-19 vaccine hesitancy and the results of the multinomial logistic regression. A total of 181 participants were used for this data analysis. Overall, 63.5% (n=115) of participants were found to be acceptant of the COVID-19 vaccine and 36.5% (n=66) were hesitant.

The hesitancy rates for majority of the predictors were lower than those who were acceptant of the vaccine (Fig. 1). For the factors “COVID-19 is not serious,” “Concerned the vaccine not effective,” and “Not knowledgeable on the vaccine,” the percentage of hesitant individuals were greater in these categories. Although 95% of the respondents deemed COVID-19 as serious, 36.5% of individuals remained hesitant towards vaccination. Most (90.0%) participants were knowledgeable about the vaccine and the remaining (n=18, 10.0%) admitted to having little knowledge of the vaccine.

**Fig 1.**
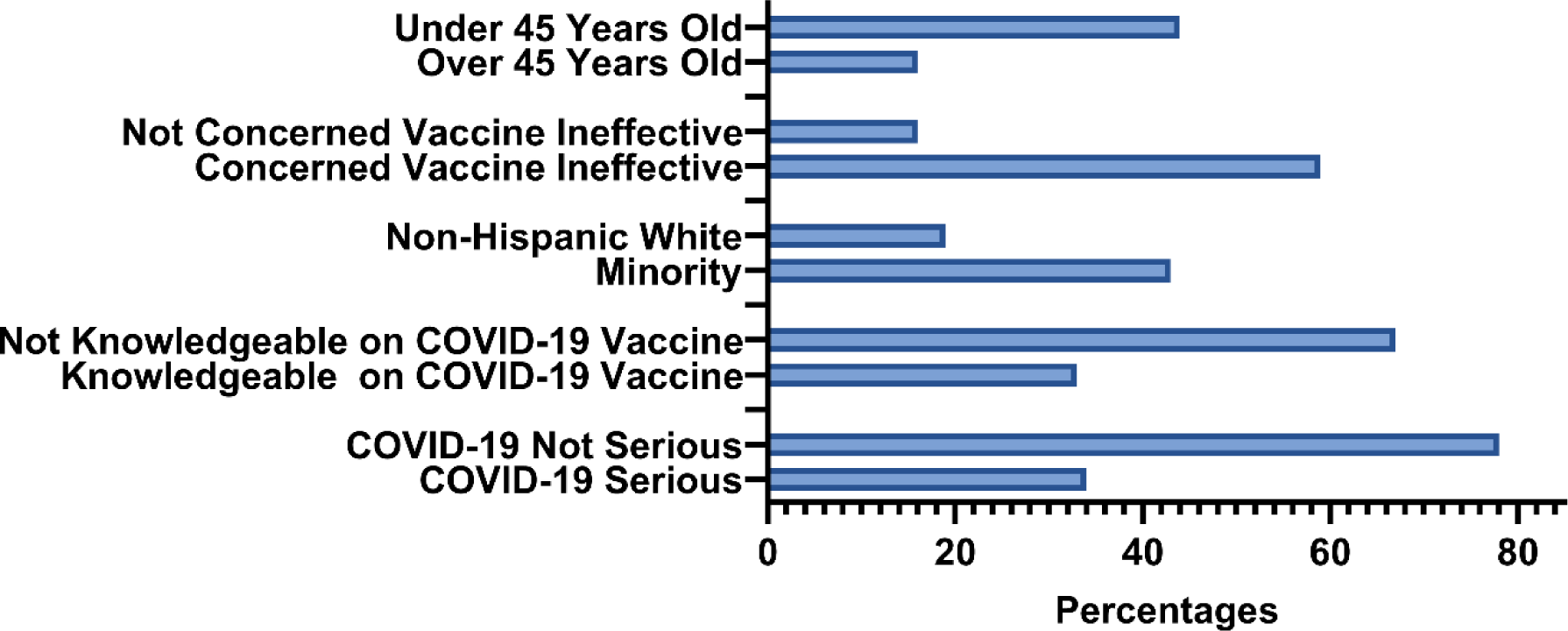
Percent of hesitant individuals towards the COVID-19 vaccine in Eastern Pennsylvania for each factor found to be significant during bivariate analysis (N = 181).

We found that participants over 45 were less likely to be hesitant towards the vaccine (OR: 0.210, 95% CI: 0.079, 0.559) than those under 45. Individuals who agreed that they were concerned that the vaccine is ineffective showed 58.6% hesitancy (OR: 8.29, 95%CI: 3.78,18.2) and 41.4% were acceptant of the vaccine. Minorities showed 43.4% hesitancy (OR: 2.77, 95% CI: 1.13, 6.79) and 56.6% acceptance of the vaccine, while non-Hispanic whites (NHW) showed 19.2% hesitancy and 80.8% acceptance. Of those who were knowledgeable about the vaccine, 66.9% expressed acceptance and 33.1% were hesitant (OR: 0.366, 95% CI: 0.108,1.25). Knowledgeability about the vaccine was found to be a significant predictor of vaccine hesitancy during the bivariate analysis, but not statistically significant according to the multivariate regression model. For those who believed the COVID-19 disease is not serious, 22.2% were acceptant of the vaccine, while 77.8% were hesitant (OR: 8.28, 95% CI: 1.11, 61.8). Figure 2 shows the odds of hesitancy for these four statistically significant predictors.

**Fig 2.**
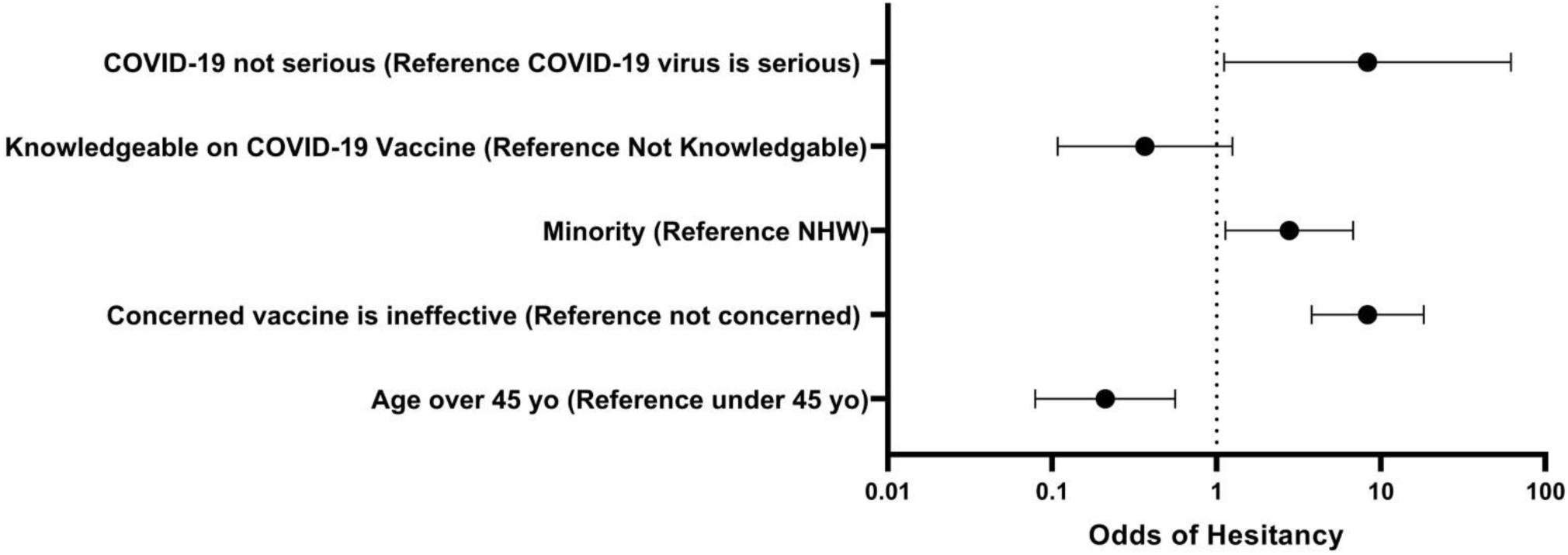
Forest plot of influential factors associated with vaccine hesitancy (Table S2). N = 181. Individuals who thought COVID-19 was serious, identified as a minority and were under 45 years of age show increased likelihood of being hesitant toward the COVID-19 vaccine. NHW – non-Hispanic white Eastern PA sample.

The top reasons selected for not receiving the COVID-19 vaccine were “I don’t think the vaccine is safe (29.6%), “Scientists do not know enough about the vaccine” (28.2%), and “The history of medical research scares me” (15.5%). Less than one in twenty unvaccinated respondents reported “I am afraid of needles” (4.2%) or “It’s against my religious beliefs” (4.2%).

The top reasons selected for receiving the COVID-19 vaccine were “I feel the vaccine is effective in preventing COVID-19” (35.1%), “I don’t want to risk transmitting COVID-19 to anyone else” (26.3%), and “I feel the vaccine is safe” (24.6%). Less than one-tenth chose “My workplace made it mandatory” (7.9%).

## Discussion

COVID-19 vaccines have shown clear efficacy in overcoming the global virus in reducing the number of hospitalizations and death from severe disease, and therefore vaccine acceptance by all will be the cornerstone of public health intervention [14]. Our focus on assessing the profile of unvaccinated individuals in our community in eastern Pennsylvania is essential to the emergence of COVID-19 vaccine acceptance. Given that disproportionate numbers of Black and Latinx communities are unvaccinated, we sought to assess the barriers to vaccine acceptance within these communities. We analyzed a convenience sample of 181 respondents amongst various areas of Eastern PA. Our findings from the summer of 2021 revealed only moderate COVID-19 vaccine acceptance (63.5%). The remaining 36.5% represent the respondents who identified as vaccine hesitant. In other published literature, vaccine hesitancy accounted for 26.3% of adult Americans across a final pool of 107,841 participants from 13 studies [13].

Although over three-fifths showed vaccine acceptance, there is still a significant amount of vaccine hesitancy that needs to be addressed. Our results indicate that the following factors were significant amongst the vaccine-hesitant: concerned vaccine not effective, not knowledgeable about COVID-19 vaccine, and COVID-19 illness is not serious. Other significant factors included age and race. Our findings in contributors to vaccine hesitancy were similar to other studies [9, 13], including concern about COVID-19 vaccine efficacy and education. Other predictors in the profile of hesitant individuals include female gender, larger household size, those with children at home, and political party affiliation [9,13].

Contrary to many other studies [7, 9,10,13] although see [15], the results of our binary regression comparing education to vaccine hesitancy suggest that education level had no impact on hesitancy. The results potentially suggest that other factors contributed to our sample’s vaccine acceptance or hesitance. Similar to other published literature [7, 9,10,13], our findings identified vaccine hesitancy being greater amongst minority populations. These results suggest that public health strategies to promote vaccine acceptance should prioritize and focus on promoting COVID-19 vaccine equity in minority groups.

Consistent with the results from previously completed investigations, vaccine acceptance reveals a correlation with knowledge about the COVID-19 vaccine [10]. In our data, those who reported not being knowledgeable about the COVID-19 vaccine were also vaccine-hesitant. In contrast, of those who are acceptant of the COVID-19 vaccine, 66.9% reported being knowledgeable about the vaccine. These findings can result from a lack of information concerning the COVID-19 vaccine or be due to the sources of the information relied upon. Knowledge about the COVID-19 vaccine intersected with trust reporting high confidence levels in health care professionals [14]. Trust and reliability of sources from an individual’s perspective can explain the vaccine hesitancy in minority populations. Whether or not an individual trusts the source of information can influence how receptive one is to information. Rapid development and information changes regarding COVID-19 and its vaccine were found to have contributed to individuals’ mistrust of outsourced information [8]. Being among the first groups to receive the vaccine when side effects and adverse reactions are least known leaves room for concern. As vaccine uptake increases, more information also arises concerning COVID-19 and FDA approved vaccinations. This may suggest that more research is needed regarding improved ways to disseminate correct and relevant information about the COVID-19 vaccines with a broader range of information, including advantages, disadvantages, and short and long-term adverse effects.

Local news and public health resources (e.g. the Centers for Disease Control and Prevention) are various ways the general population receives information concerning COVID-19 and its vaccine [7, 10]. The following recommendations are offered to make information concerning COVID-19 vaccines better received and promote vaccine acceptance. A focus needs to be on minority populations as they are the most vaccine-hesitant [13]. Community-based talks, direct personal contact, and realistic outcomes can be used to encourage those who are hesitant about vaccine uptake. Contrary to the inaccurate information being disseminated across platforms [7], accurate information that is respectfully delivered by trusted messengers must take its place. Studies have concluded that media and scientific outlets have begun to release accurate information to enlighten the general public about the COVID-19 vaccine to mitigate vaccine hesitancy [9]. Informational sources concerning COVID-19 and its vaccine should be explicit and clear. Information needs to be supported with medical evidence that can be easily understood, championed by trusted experts and trusted members or leaders within the communities of interest. Implementing inclusive programs for informational purposes, including live seminars and enlightening community groups, can alleviate those hesitant and promote acceptance [8].

Although this investigation involved responding quickly to a dynamic public health situation, the observed results are limited by only a moderate sample size and occasional missing responses on select variables. Data collection through self-report reflects another limitation as self-assessment of COVID-19 knowledge can introduce biases. Another limitation was access to unvaccinated individuals. Most of the population surveyed had already received the vaccine or were receptive to getting it. We can not discount that some participants holding strong anti-vaccine beliefs [15] were less likely to participate in this voluntary study. Another limitation was the few native Spanish-speaking participants in our sample. Future studies should seek to include sizable proportions of unvaccinated individuals [15] along with the addition of bilingual research assistants and interpreters to their team. Future investigations that include an appreciable number of unvaccinated participants may further clarify the overall profile of unvaccinated minorities, allowing for the development of a more nuanced strategy to address their concerns and encourage vaccine and booster acceptance among all ages [19].

## Conclusion

The objectives of this study were to characterize what variables affect COVID-19 vaccination hesitancy in Black and Latinx communities in Eastern Pennsylvania. The variables which had an impact on vaccination hesitancy were younger age, concern about vaccine effectiveness, race, knowledge about the COVID-19 vaccine, and perceived severity of COVID-19 disease. This investigation enables better targeted outreach to unvaccinated individuals within the minority communities. This data would help public health researchers understand why these minorities are more hesitant to receive the COVID-19 vaccine. From there, they will be able to provide data to participants that shows the pros of receiving the vaccination, easing their hesitancy. Overall, these findings will help contribute to eliminating disparities and ending the COVID-19 pandemic.

## Supporting information

educational COVID-19 materials

Survey in English

## Data Availability

Raw data is available as a supplemental file.

## Acknowledgements

This research was supported by the Health Resources Services Administration (D34HP31025). Dr. Reema Persad-Clem PhD, MPH, Elizabeth Kuchinski BS, MPH, Lavinia Harrison, Jonique Depina, MBS for there are thanked for their guidance and feedback throughout the writing process.

