## Supplementary material for "Assessing the Profile of Unvaccinated COVID-19 Individuals in African American and Latinx Communities in Eastern Pennsylvania": educational COVID-19 materials

### Education Info about COVID-19

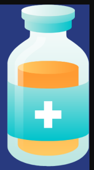

**01** The **Moderna** and **Pfizer vaccines** are both **mRNA** vaccines which contain mRNA codes for **spike proteins** inside of a **protective capsule** which allows it to enter the cell. These require **2 shots** to be fully effective.

**02** The **Johnson and Johnson vaccine** is a **non-replicative adenovirus** with spike protein DNA. A weakened virus is produced and used to carry the spike protein DNA into the cells.This vaccine requires **one shot**.

**03** Both of these vaccines deliver instructions to our cells, but the instructions do not enter the **nucleus of the cell**, where **DNA** is kept. The vaccine is injected in the **muscle** in the **upper arm** and does not travel anywhere else in the body.

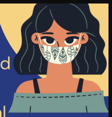

**04** **Immunizations** train the body to build **IGG's (antibodies)** before they are needed in order to make an individual ready to fight that **illness**.

**05** Although you can still get **COVID-19** if you have been vaccinated, vaccination significantly **reduces transmission**, **hospitalization**, and **death related to COVID-19**.

**06** The vaccines may cause certain side effects or reactions such as: **reactogenicity** which is an **inflammatory response** which could be **local** or **systemic** and last a few days, or **immunogenicity** which is a **normal protective response** from the **immune system**.

**07** The most **common of these side effects** can be seen as arm soreness, tiredness, headache, muscle pains, chills, fever, and nausea. Taking **Tylenol or Ibuprofen** will ease these side effects after vaccination.

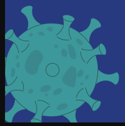

**08** The Johnson and Johnson vaccine has a rare but increased risk of **Thrombocytopenia syndrome** for women aged 15-50. This is a **clotting disorder** where major **vessels** in the body can become blocked. The chance of this reaction is currently around **7 out of 1 million vaccinations.**

**09** The **Moderna vaccine** has an **efficacy rate of 94.1%**, the **Pfizer vaccine** has an **efficacy rate of 95%**, and the **Johnson and Johnson vaccine** has an **efficacy rate of 72%**. Efficacy is determined by how many individuals get the virus, even after being vaccinated, the **higher the efficacy rate the less individuals that got the virus.**

**10** If you need help finding or accessing resources in order to get the vaccine, **scan the QR code below, or follow the link!**

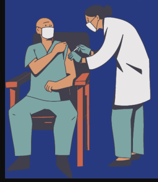

vaccines.gov

SCAN ME
