## Supplementary material for "Assessing the Profile of Unvaccinated COVID-19 Individuals in African American and Latinx Communities in Eastern Pennsylvania": Survey in English

Your Account Expires in 9 Days
Keep access to your survey data and paid features.
Enable auto-renew

### COVID-19 Survey Questions



**Your feedback is important to us**

Thank you! We read every response. Privacy notice

Do the analyze visualizations and tools help you gather data insights from your survey?

- Yes
- No

Continue

Integrate SurveyMonkey with tools you already use, like **Power BI**, **Tableau**, **Zoom**, and **Salesforce**, to automate workflows, create deeper insights, and get more value out of your work day.
Get a Demo

RESPONDENTS: 198 of 198

ADD TO DASHBOARD SAVE AS

QUESTION SUMMARIES | INSIGHTS AND DATA TRENDS | INDIVIDUAL RESPONSES

All Pages

Page 1: Demographic Questions

**Q1**

Save as

#### County of Residence

Answered: 192 Skipped: 6

RESPONSES (192) | WORD CLOUD | TAGS (0)

Filter: by tag

Search responses

Showing **192** responses

Bucks county
7/22/2021 10:16 PM View respondent's answers Add tags

United States
7/22/2021 9:56 PM View respondent's answers Add tags

Lackawanna
7/21/2021 2:52 PM View respondent's answers Add tags

Hazel Township
7/21/2021 1:14 PM View respondent's answers Add tags

Oops, Something went wrong

**Q2**

Customize Save as

#### Gender: Pick one

Answered: 198 Skipped: 0

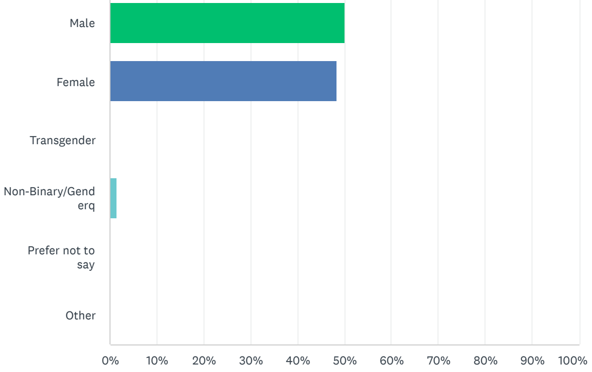


| ANSWER CHOICES | RESPONSES | |
|---|---|---|
| Male | 50.00% | 99 |
| Female | 48.48% | 96 |
| Transgender | 0.00% | 0 |
| Non-Binary/Genderq | 1.52% | 3 |
| Prefer not to say | 0.00% | 0 |
| Other | 0.00% | 0 |
| TOTAL | | 198 |

Page 2: Demographic Questions

Q3

Save as

#### Explain your answer to question 2

Answered: 0 Skipped: 198

RESPONSES (0) WORD CLOUD TAGS (0)


NEW!

**Introducing Sentiment Analysis**

Detect the feeling and sentiment behind written responses.

Watch a demo

Try it!

Upgrade


Filter: by tag

Search responses

Showing **0** responses


#### Your feedback is important to us

Thank you! We read every response. Privacy notice

Do the analyze visualizations and tools help you gather data insights from your survey?

Yes

No

Continue


Oops, Something went wrong

Your feedback is important to us

Thank you! We read every response. Privacy notice

Do the analyze visualizations and tools help you gather data insights from your survey?

Yes

No

Continue

Page 3: Demographic Questions

Q4

Customize Save as

#### Race/Ethnicity: Select all that apply to you.

Answered: 194 Skipped: 4

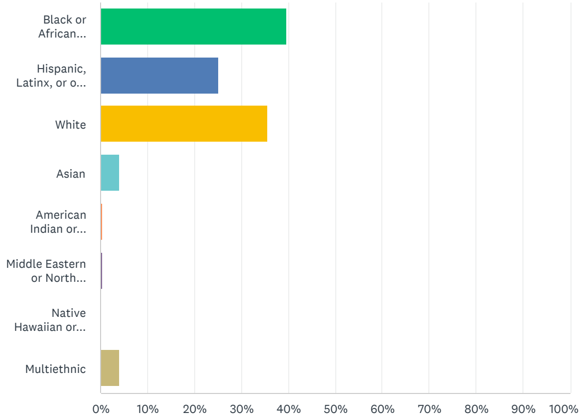


| ANSWER CHOICES | RESPONSES | |
|---|---|---|
| Black or African American | 39.69% | 77 |
| Hispanic, Latinx, or of Spanish Origin | 25.26% | 49 |
| White | 35.57% | 69 |
| Asian | 4.12% | 8 |
| American Indian or Alaska native | 0.52% | 1 |
| Middle Eastern or North African | 0.52% | 1 |
| Native Hawaiian or other Pacific Islander | 0.00% | 0 |
| Multiethnic | 4.12% | 8 |
| Total Respondents: 194 | | |

Q5

Customize Save as

#### Age in Years: Pick one

Answered: 195 Skipped: 3

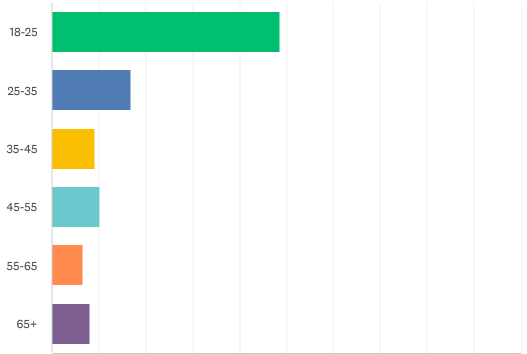


Oops, Something went wrong

0% 10% 20% 30% 40% 50% 60% 70% 80% 90% 100%

| ANSWER CHOICES | RESPONSES | |
|---|---|---|
| 18-25 | 48.72% | 95 |
| 25-35 | 16.92% | 33 |
| 35-45 | 9.23% | 18 |
| 45-55 | 10.26% | 20 |
| 55-65 | 6.67% | 13 |
| 65+ | 8.21% | 16 |
| TOTAL | | 195 |


#### Your feedback is important to us

Thank you! We read every response. Privacy notice

Do the analyze visualizations and tools help you gather data insights from your survey?

- Yes
- No

Continue


## Q6

Customize Save as

##### What is your highest level of education?

Answered: 192 Skipped: 6

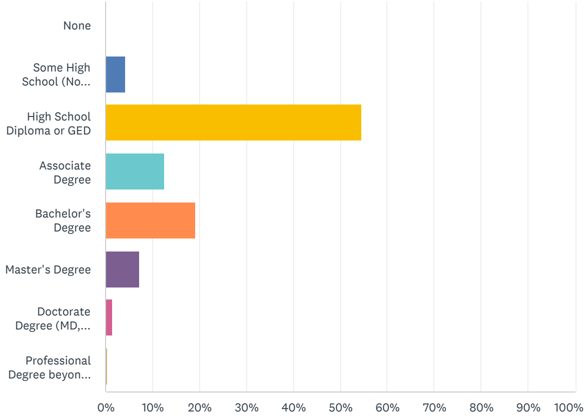


| ANSWER CHOICES | RESPONSES | |
|---|---|---|
| None | 0.00% | 0 |
| Some High School (No Diploma) | 4.17% | 8 |
| High School Diploma or GED | 54.69% | 105 |
| Associate Degree | 12.50% | 24 |
| Bachelor's Degree | 19.27% | 37 |
| Master's Degree | 7.29% | 14 |
| Doctorate Degree (MD, PhD, etc.) | 1.56% | 3 |
| Professional Degree beyond Bachelor’s | 0.52% | 1 |
| TOTAL | | 192 |


Oops, Something went wrong


Page 4: COVID-19 Questions

## Q7

Customize Save as

##### Have you ever had COVID-19?

Answered: 194 Skipped: 4

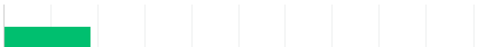

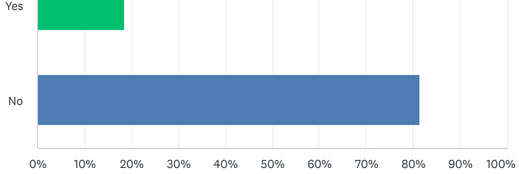


| ANSWER CHOICES | RESPONSES | |
|---|---|---|
| Yes | 18.56% | 36 |
| No | 81.44% | 158 |
| TOTAL | | 194 |


Your feedback is important to us

Thank you! We read every response. Privacy notice

Do the analyze visualizations and tools help you gather data insights from your survey?

Yes

No

Continue


Page 5: COVID-19 Questions

Q8

Customize Save as

#### If yes, how worried are you about getting it agian?

Answered: 36 Skipped: 162

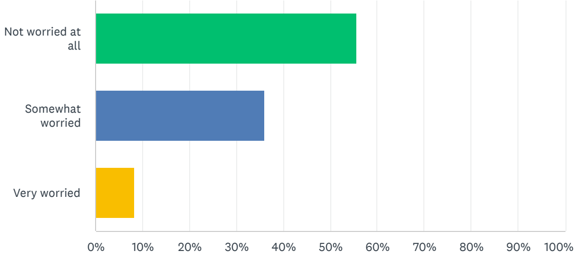


| ANSWER CHOICES | RESPONSES | |
|---|---|---|
| Not worried at all | 55.56% | 20 |
| Somewhat worried | 36.11% | 13 |
| Very worried | 8.33% | 3 |
| TOTAL | | 36 |

Page 6: COVID-19 Questions

Q9

Customize Save as

#### If no, how worried are you about getting it?

Answered: 153 Skipped: 45

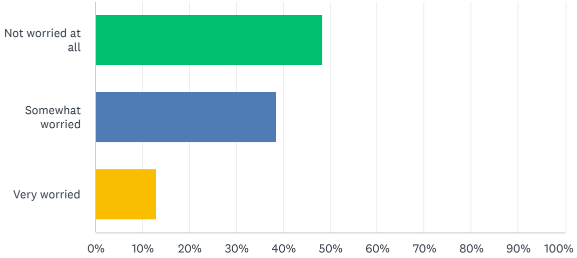



Oops, Something went wrong

| ANSWER CHOICES | RESPONSES | |
|---|---|---|
| Not worried at all | 48.37% | 74 |
| Somewhat worried | 38.56% | 59 |
| Very worried | 13.07% | 20 |
| TOTAL | | 153 |

Page 7: COVID-19 Questions

## Q10

Customize Save as

##### Do you have paid sick leave in case you get sick?

Answered: 189 Skipped: 9

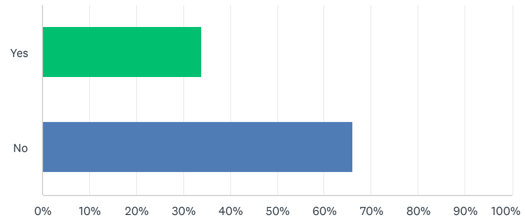


| ANSWER CHOICES | RESPONSES | |
|---|---|---|
| Yes | 33.86% | 64 |
| No | 66.14% | 125 |
| TOTAL | | 189 |

## Q11

Customize Save as

##### Do you have health insurance or Medicaid/Medicare in case you get sick?

Answered: 190 Skipped: 8

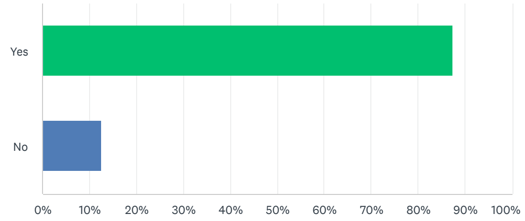


| ANSWER CHOICES | RESPONSES | |
|---|---|---|
| Yes | 87.37% | 166 |
| No | 12.63% | 24 |
| TOTAL | | 190 |

## Q12

Customize Save as

Your feedback is important to us
Thank you! We read every response. Privacy notice
Do the analyze visualizations and tools help you gather data insights from your survey?
Yes
No
Continue
Oops, Something went wrong

#### To what extent would you say that you have been affected by COVID-19?

Answered: 190 Skipped: 8

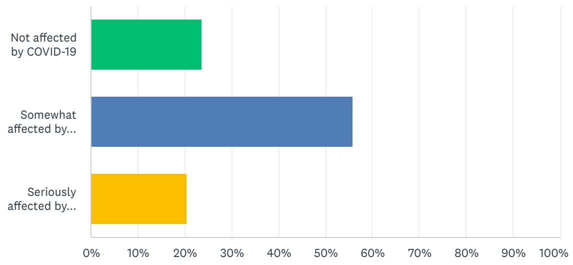


| ANSWER CHOICES | RESPONSES | |
|---|---|---|
| Not affected by COVID-19 | 23.68% | 45 |
| Somewhat affected by COVID-19 | 55.79% | 106 |
| Seriously affected by COVID-19 | 20.53% | 39 |
| TOTAL | | 190 |


Your feedback is important to us

Thank you! We read every response. Privacy notice

Do the analyze visualizations and tools help you gather data insights from your survey?

Yes

No

Continue


Q13 Customize Save as

#### How serious do you think the COVID-19 virus is?

Answered: 190 Skipped: 8

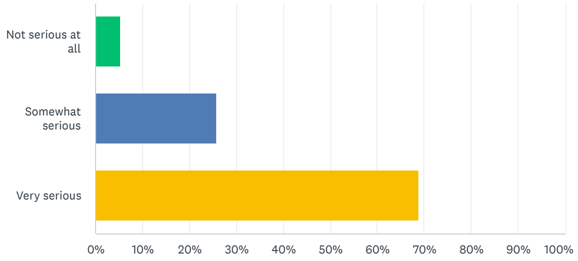


| ANSWER CHOICES | RESPONSES | |
|---|---|---|
| Not serious at all | 5.26% | 10 |
| Somewhat serious | 25.79% | 49 |
| Very serious | 68.95% | 131 |
| TOTAL | | 190 |

Q14 Customize Save as


Oops, Something went wrong


#### Where do you obtain your information on COVID-19?

Answered: 190 Skipped: 8

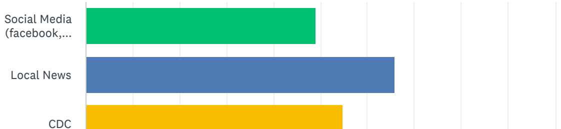

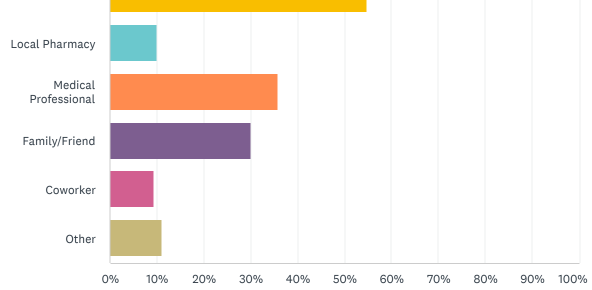


| ANSWER CHOICES | RESPONSES | |
|---|---|---|
| Social Media (facebook, Snapchat, Instagram, TikTok, Twitter) | 48.95% | 93 |
| Local News | 65.79% | 125 |
| CDC | 54.74% | 104 |
| Local Pharmacy | 10.00% | 19 |
| Medical Professional | 35.79% | 68 |
| Family/Friend | 30.00% | 57 |
| Coworker | 9.47% | 18 |
| Other | 11.05% | 21 |
| **Total Respondents: 190** | | |

Your feedback is important to us

Thank you! We read every response. Privacy notice

Do the analyze visualizations and tools help you gather data insights from your survey?

Yes

No

Continue

Page 8: COVID-19 Questions

Q15

Save as

#### Explain your answer to question 14

Answered: 11 Skipped: 187

RESPONSES (11) WORD CLOUD TAGS (0)

Filter: by tag

Search responses

Showing **11** responses

Search multiple search engines and sources for information that can be validated

7/20/2021 9:28 AM View respondent's answers Add tags

Other

7/19/2021 12:53 PM View respondent's answers Add tags

The internet mostly via the news filter on google

7/19/2021 11:24 AM View respondent's answers Add tags

I would read articles and do some research myself

7/19/2021 9:50 AM View respondent's answers Add tags

Oops, Something went wrong

Page 9: COVID-19 Questions

Q16

Customize Save as

What type of impact do you believe these sources (listed above) have had on

What type of impact do you believe these sources (listed above) have had on your community?

Answered: 184 Skipped: 14

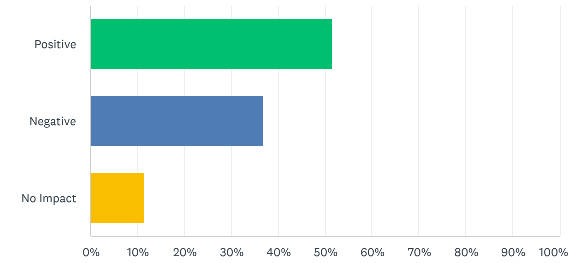


| ANSWER CHOICES | RESPONSES | |
|---|---|---|
| Positive | 51.63% | 95 |
| Negative | 36.96% | 68 |
| No Impact | 11.41% | 21 |
| TOTAL | | 184 |


Your feedback is important to us

Thank you! We read every response. Privacy notice

Do the analyze visualizations and tools help you gather data insights from your survey?

Yes

No

Continue


Q17

Customize Save as

Personally, what stage do you think we are in the timeline of the COVID-19 pandemic?

Answered: 183 Skipped: 15

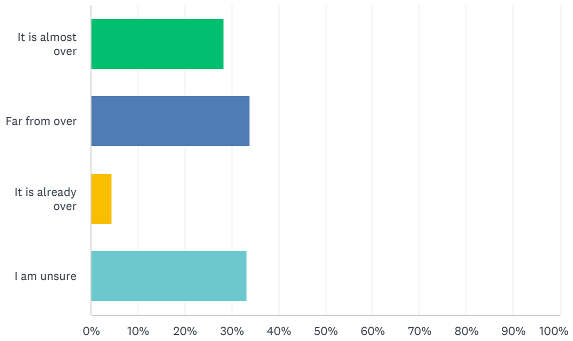


| ANSWER CHOICES | RESPONSES | |
|---|---|---|
| It is almost over | 28.42% | 52 |
| Far from over | 33.88% | 62 |
| It is already over | 4.37% | 8 |
| I am unsure | 33.33% | 61 |
| TOTAL | | 183 |


Oops, Something went wrong


#### Page 10: COVID-19 Vaccine Questions

Q18

Customize Save as

How knowledgeable would you say you are about the COVID-19 vaccine?

Answered: 184 Skipped: 14

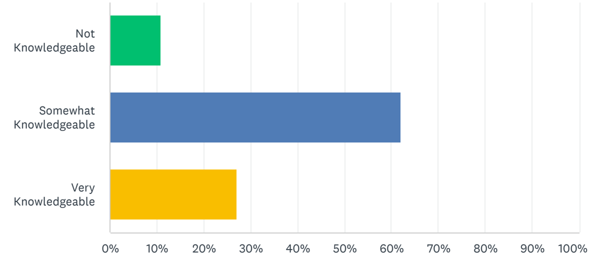


| ANSWER CHOICES | RESPONSES | |
|---|---|---|
| Not Knowledgeable | 10.87% | 20 |
| Somewhat Knowledgeable | 61.96% | 114 |
| Very Knowledgeable | 27.17% | 50 |
| TOTAL | | 184 |

Your feedback is important to us

Thank you! We read every response. Privacy notice

Do the analyze visualizations and tools help you gather data insights from your survey?

Yes

No

Continue

## Q19

Customize Save as

Where do you get most of your information about the vaccine?

Answered: 182 Skipped: 16

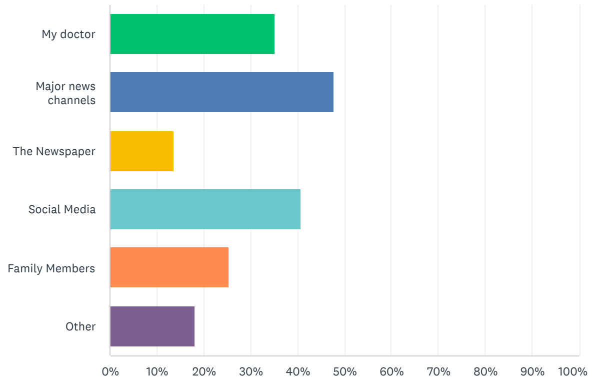


| ANSWER CHOICES | RESPONSES | |
|---|---|---|
| My doctor | 35.16% | 64 |
| Major news channels | 47.80% | 87 |
| The Newspaper | 13.74% | 25 |
| Social Media | 40.66% | 74 |
| Family Members | 25.27% | 46 |
| Other | 18.13% | 33 |
| Total Respondents: 182 | | |

Oops, Something went wrong

Page 11: COVID-19 Vaccine Questions

## Q20

Save as

Explain your answer to question 19

Answered: 22   Skipped: 176

RESPONSES (22)   WORD CLOUD   TAGS (0)

Filter: by tag

Search responses

Showing **22** responses

I have done my own outside research and applied my previous knowledge of vaccines in general

7/22/2021 9:59 PM   View respondent's answers   Add tags

CDC

7/21/2021 6:20 AM   View respondent's answers   Add tags

Medical journals, CDC site

7/20/2021 10:04 PM   View respondent's answers   Add tags

See answer 13/14

7/20/2021 9:30 AM   View respondent's answers   Add tags


#### Your feedback is important to us

Thank you! We read every response. Privacy notice

Do the analyze visualizations and tools help you gather data insights from your survey?

- Yes
- No

Continue


Page 12: COVID-19 Vaccine Questions

**Q21**   Customize   Save as

#### I have concerns that the vaccine is not effective

Answered: 181   Skipped: 17

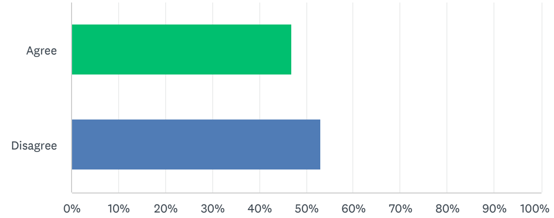


| ANSWER CHOICES | RESPONSES | |
|---|---|---|
| Agree | 46.96% | 85 |
| Disagree | 53.04% | 96 |
| TOTAL | | 181 |

**Q22**   Customize   Save as

#### COVID vaccine is now available to protect people from getting the infection. If you had the opportunity to get the vaccine, would you...

Answered: 181   Skipped: 17

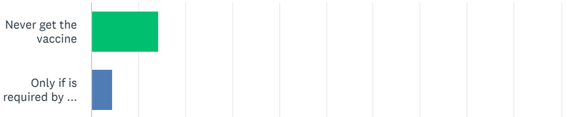


Oops, Something went wrong

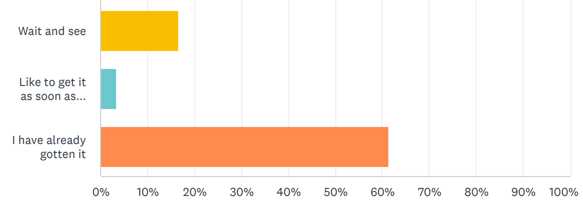


| ANSWER CHOICES | RESPONSES | |
|---|---|---|
| Never get the vaccine | 14.36% | 26 |
| Only if is required by the state or the employer | 4.42% | 8 |
| Wait and see | 16.57% | 30 |
| Like to get it as soon as possible | 3.31% | 6 |
| I have already gotten it | 61.33% | 111 |
| TOTAL | | 181 |

Your feedback is important to us

Thank you! We read every response. Privacy notice

Do the analyze visualizations and tools help you gather data insights from your survey?

Yes

No

Continue

Page 13: COVID-19 Vaccine Questions

## Q23

Customize Save as

##### If you have not yet gotten the vaccine, how safe do you think the vaccine is?

Answered: 70 Skipped: 128

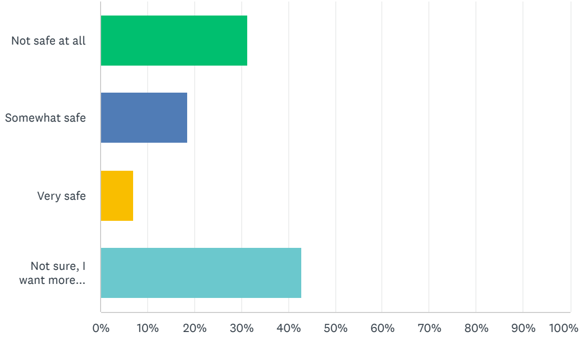


| ANSWER CHOICES | RESPONSES | |
|---|---|---|
| Not safe at all | 31.43% | 22 |
| Somewhat safe | 18.57% | 13 |
| Very safe | 7.14% | 5 |
| Not sure, I want more information | 42.86% | 30 |
| TOTAL | | 70 |

Oops, Something went wrong

Page 14: COVID-19 Vaccine Questions

## Q24

Customize Save as

##### Which of the following best explains why you chose to be vaccinated?

Answered: 111 Skipped: 87

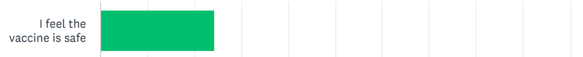

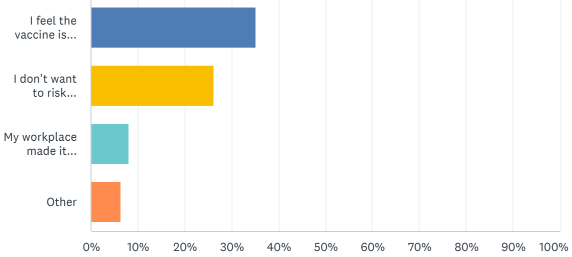


| ANSWER CHOICES | RESPONSES | |
|---|---|---|
| I feel the vaccine is safe | 24.32% | 27 |
| I feel the vaccine is effective in preventing COVID | 35.14% | 39 |
| I don't want to risk transmitting COVID-19 to anyone else | 26.13% | 29 |
| My workplace made it mandatory | 8.11% | 9 |
| Other | 6.31% | 7 |
| TOTAL | | 111 |


#### Your feedback is important to us

Thank you! We read every response. Privacy notice

Do the analyze visualizations and tools help you gather data insights from your survey?

Yes

No

Continue


Page 15: COVID-19 Vaccine Questions

**Q25**

Save as

#### Explain your answer to question 24

Answered: 7 Skipped: 191

RESPONSES (7) WORD CLOUD TAGS (0)

Filter: by tag

Search responses

Showing **7** responses

Preexisting medical issues that result in compromised immune system

7/20/2021 9:31 AM View respondent's answers Add tags

Travel, personal health reasons, to avoid issues with work, school, etc.

7/19/2021 2:26 PM View respondent's answers Add tags

Loved ones

7/19/2021 10:59 AM View respondent's answers Add tags

Jail made it

7/16/2021 10:09 AM View respondent's answers Add tags

Oops, Something went wrong

Page 16: COVID-19 Vaccine Questions

**Q26**

Customize Save as

#### If you do not want to get the vaccine, which answer best describes your reason for not getting the vaccine?

Answered: 69 Skipped: 129

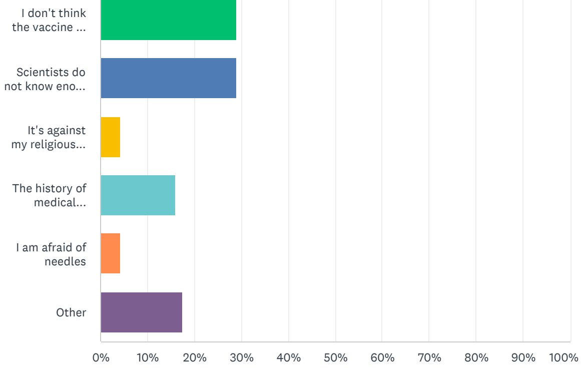


| ANSWER CHOICES | RESPONSES | |
|---|---|---|
| I don't think the vaccine is safe | 28.99% | 20 |
| Scientists do not know enough about the vaccine | 28.99% | 20 |
| It's against my religious beliefs | 4.35% | 3 |
| The history of medical research scares me | 15.94% | 11 |
| I am afraid of needles | 4.35% | 3 |
| Other | 17.39% | 12 |
| TOTAL | | 69 |

Page 17: COVID-19 Vaccine Questions

**Q27**

Save as

#### Explain your answer to question 26

Answered: 10 Skipped: 188

RESPONSES (10) WORD CLOUD TAGS (0)

Filter: by tag

Search responses

Showing **10** responses

The side effects that I have heard about concern me

7/20/2021 9:14 AM View respondent's answers Add tags

My heritage

7/19/2021 1:22 PM View respondent's answers Add tags

I want to get it, just don't got time I wanna get the johnson

7/19/2021 11:15 AM View respondent's answers Add tags

I don't think it's safe

7/19/2021 10:58 AM View respondent's answers Add tags

Page 18: COVID-19 Vaccine Questions

**Q28**

Customize Save as


Your feedback is important to us

Thank you! We read every response. Privacy notice

Do the analyze visualizations and tools help you gather data insights from your survey?

Yes

No

Continue

Oops, Something went wrong

If you want the vaccine, do you know where to get the vaccine?

Answered: 178 Skipped: 20

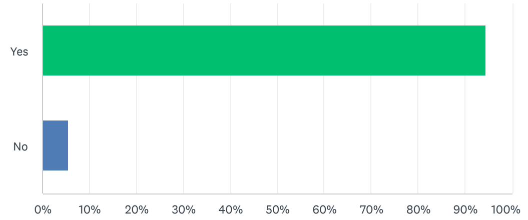


| ANSWER CHOICES | RESPONSES | |
|---|---|---|
| Yes | 94.38% | 168 |
| No | 5.62% | 10 |
| TOTAL | | 178 |


#### Your feedback is important to us

Thank you! We read every response. Privacy notice

Do the analyze visualizations and tools help you gather data insights from your survey?

Yes

No

Continue

ENGLISH

About SurveyMonkey • Careers • Developers • Privacy Notice • California Privacy Notice • Email Opt-In • Help • Cookies Notice • Do Not Sell My Personal Information

Copyright © 1999-2022 SurveyMonkey

ACCREDITED BUSINESS

TrustedSite CERTIFIED SECURE
